# Determinants of self-reported symptoms and testing for COVID in Canada using a nationally representative survey

**DOI:** 10.1101/2020.05.29.20109090

**Authors:** Daphne C Wu, Prabhat Jha, Teresa Lam, Patrick Brown, Hellen Gelband, Nico Nagelkerke, H. Chaim Birnboim, Angus Reid, on behalf of the Action to Beat Coronavirus in Canada/Action pour Battre le Coronavirus (**Ab-C**) Study Group

## Abstract

In April 2020, the first-ever nationally representative survey in Canada polled 4,240 adults age 18 years and older about their COVID experience in March, early in the epidemic. We examined determinants of COVID symptoms, defined as fever plus difficulty breathing/shortness of breath, dry cough so severe that it disrupts sleep, and/or loss of sense of smell; and testing for SARS-CoV-2 by respondents and/or household members. About 8% of Canadians reported that they and/or one or more household members experienced COVID symptoms. Symptoms were more common in younger than older adults, and among visible minorities. Overall, only 3% of respondents and/or household members reported testing for SARS-CoV-2. Being tested was associated with having COVID symptoms, Indigenous identity, and living in Quebec. Periodic nationally representative surveys—including high-risk older populations—of symptoms, as well as SARS-CoV-2 antibodies, are required in many countries to understand the pandemic and prepare for the future.

## Introduction

The pandemic of SARS-CoV-2 infection causing coronavirus disease-2019 (COVID-19, hereafter “COVID”) has affected Canada and many other high-income countries (Eggertson and Wolfville, 2020). In these settings where reliable data can be gathered, a combination of population-based surveys (including surveys and testing), hospitalizations, and mortality data can produce an accurate profile of the impact of the epidemic.

We report on the results of the first nationally-representative poll of self-reported COVID symptoms conducted in Canada by the Angus Reid Forum in early April 2020 covering symptoms reported mostly in March 2020, prior to the peak month of cases in April. Our goal was to understand the distribution and determinants of Canadians reporting possible COVID symptoms. We also sought to understand who underwent testing for SARS-CoV-2 using the current standard (PCR-based) test. We discuss these findings in the context of the age distribution of COVID hospitalizations and deaths, and a planned survey of antibodies to SARS-CoV-2 in a random sample of Canadians.

## Results

To determine the representativeness of the respondents, we compared their socio-demographic characteristics to those of the Canadian population in 2019 (Table 1). Overall, the survey respondents were broadly representative of Canadian society in terms of gender, age, regional distribution, and numbers of household members. Survey respondents were less representative of Canada in terms of ethnicities other than Indigenous. The survey had fewer single-member households than in the Canadian census, and had slightly higher education levels than did the 2019 Canadian population. Approximately 0.3 million (out a total population of 38 million) Canadians are estimated to live in nursing homes/long-term care institutions (Canadian Life and Health Insurance Association, 2014). This would represent less than 1% of the expected population in the survey (or about 40 participants). However, such populations were under-represented in the survey.

**Table 1.** Socio-demographic characteristics of respondents (N=4,240) as compared to the Canadian population in 2019.

| Characteristics | Completed survey,<br>sample size |  | Canadian<br>population,<br>2019 (%)<br>(Statistics<br>Canada, 2016;<br>Statistics<br>Canada, 2020a) |
| --- | --- | --- | --- |
|  | n | % |  |
| **Gender** |  |  |  |
| Male | 2017 | 47.9 | 49.4 |
| Female | 2205 | 52.0 | 50.6 |
| Other | 18 | 0.4 |  |
| **Age group** |  |  |  |
| 18-45 years | 2007 | 47.3 | 46.4 |
| 46-65 years | 1511 | 35.6 | 33.4 |
| 66+ years | 722 | 17.0 | 20.2 |
| **Education** |  |  |  |
| High school and under | 1064 | 25.1 | 35.2 |
| Some college/university and higher | 3176 | 74.9 | 64.8 |
| **Visible minority** |  |  |  |
| No | 3686 | 86.9 | 71.4 |
| Yes | 554 | 13.1 | 28.7 |
| **Number of household members** |  |  |  |
| Lived alone | 672 | 15.9 | 28.2 |
| Two people | 1615 | 38.1 | 34.4 |
| Three people | 804 | 19.0 | 15.2 |
| Four or more people | 1149 | 27.1 | 22.2 |
| **Ethnicity** |  |  |  |
| Indigenous<sup>+</sup> | 226 | 5.3 | 5.0 |
| English and other European | 3567 | 84.1 | 73.1 |
| Others | 365 | 8.6 | 22.0 |
| Rather Not Say | 82 | 1.9 |  |
| **Province/region** |  |  |  |
| Ontario | 1600 | 37.7 | 38.9 |
| British Columbia | 554 | 13.1 | 13.8 |
| Quebec | 1020 | 24.1 | 22.7 |
| Alberta | 475 | 11.2 | 11.2 |
| Manitoba | 150 | 3.5 | 3.5 |
| Saskatchewan | 133 | 3.1 | 3.0 |
| Atlantic provinces | 308 | 7.3 | 6.6 |

\*All comparisons are from the 2016 Census, except Province/region which is from 2019 Statistics Canada population data (Statistics Canada, 2020a).
<sup>+</sup>Indigenous populations including First Nations, Inuit, or Métis.

Of the 4,240 respondents, 334 (7.9% after applying survey weights) reported COVID symptoms, defined as the respondent reporting himself/herself and/or at least one of the household members experiencing a combination of fever (with or without hallucinations) <u>and</u> any of i) difficulty breathing/shortness of breath <u>or</u> ii) dry cough so severe that it disrupts sleep <u>or</u> iii) a loss of a sense of smell. Of these, 210 (5.0%) reported COVID symptoms only in themselves. The adjusted odds ratio (OR) of the respondent having symptoms when at least one of the household members reported symptoms was 1.45 (95% CI 1.41-1.49); the OR was similar for at least one household member having symptoms if the respondent reported symptoms. In terms of testing for SARS-CoV-2, 126 (or 3.0%) reported some household testing or being scheduled for testing, and only 68 (or 1.6%) reported that they have been or are scheduled for testing. Details of the variation in COVID symptoms and SARS-CoV-2 testing in this sample across provinces have already been published (Angus Reid Institute, 2020).

Table 2 shows the OR of respondents or a member of the household, and respondents only having COVID symptoms after adjustment for other variables examined. We excluded 99 (or 2.3%) respondents who did not report on at least one of the variables. The proportion of respondents reporting COVID symptoms within the household decreased with age: 11.2% of those aged 18-45 years, 5.6% among those aged 46-65 years, and 3.1% among those aged 66 years and older. The lower prevalence at higher ages was similar among those reporting COVID symptoms only themselves. After controlling for gender, province, age, ethnicity, visible minority, and number of household members, older adults were significantly less likely to report having COVID symptoms themselves or within the household (age 46-65, OR=0.55, 0.42-0.71; age 66+, OR=0.30, 0.18-0.47). Those who reported themselves as a visible minority were significantly more likely to have COVID symptoms (12.9%) compared to those who did not (7.1%; adjusted OR=1.55; 1.08-2.20). Similar results were found when we examined COVID symptoms only among respondents. The associations changed substantially between unadjusted and adjusted analyses, most notably for ethnicity, and somewhat for being a visible minority, suggesting that some residual confounding factors were present. Results using a narrower definition of COVID symptoms, namely having fever, difficulty breathing/shortness of breath, and severe dry cough were similar (data not shown).

**Table 2.** Logistic regression models for respondents or a member of household, and respondents only having COVID symptoms.

| Characteristics | Respondents or a member of household having symptoms |  |  | Respondents only having symptoms |  |  |
| --- | --- | --- | --- | --- | --- | --- |
|  | COVID symptoms positive/negative |  | Adjusted odds ratios (95% CI) | COVID symptoms positive/negative |  | Adjusted odds ratios (95% CI) |
|  | N=324/3817 | % |  | N=202/393<br>9 | % |  |
| **Gender** |  |  |  |  |  |  |
| Male | 149/1840 | 7.5 | Ref | 85/1904 | 4.3 | Ref |
| Female | 175/1977 | 8.1 | 1.11 (0.89-1.40) | 117/2036 | 5.4 | 1.30 (0.97-1.74) |
| **Age group** |  |  |  |  |  |  |
| 18-45 years | 218/1724 | 11.2 | Ref | 126/1816 | 6.5 | Ref |
| 46-65 years | 84/1403 | 5.6 | **0.55 (0.42-0.71)** | 61/1426 | 4.1 | **0.61 (0.44-0.84)** |
| 66+ years | 22/690 | 3.1 | **0.30 (0.18-0.47)** | 15/697 | 2.1 | **0.29 (0.16-0.50)** |
| **Education** |  |  |  |  |  |  |
| Some college/university and higher | 237/2874 | 8.4 | Ref | 150/2961 | 4.8 | Ref |
| High school and under | 87/943 | 7.6 | 1.25 (0.96-1.62) | 52/978 | 5.1 | 1.16 (0.83-1.61) |
| **Visible minority** |  |  |  |  |  |  |
| No | 256/3359 | 7.1 | Ref | 155/3460 | 4.3 | Ref |
| Yes | 68/458 | 12.9 | **1.55 (1.08-2.20)** | 47/479 | 8.9 | **2.08 (1.34-3.15)** |
| **Number of household members** |  |  |  |  |  |  |
| Two people | 108/1479 | 6.8 | Ref | 79/1507 | 5.0 | Ref |
| Lived alone | 29/626 | 4.4 | 0.69 (0.45-1.02) | 29/626 | 4.5 | 1.01 (0.64-1.53) |
| Three people | 77/704 | 9.9 | 1.17 (0.85-1.61) | 45/736 | 5.8 | 0.86 (0.57-1.28) |
| Four or more people | 110/1008 | 9.8 | 1.19 (0.89-1.59) | 48/1070 | 4.3 | 0.72 (0.49-1.05) |
| **Ethnicity** |  |  |  |  |  |  |
| English and other European | 252/3305 | 7.1 | Ref | 158/3399 | 4.4 | Ref |
| Indigenous* | 27/195 | 12.2 | 1.31 (0.83-2.01) | 19/203 | 8.5 | 1.34 (0.76-2.24) |
| Others | 45/317 | 12.4 | 1.20 (0.79-1.80) | 25/337 | 7.0 | 0.87 (0.50-1.47) |
| **Province/region** |  |  |  |  |  |  |
| Ontario | 129/1448 | 8.2 | Ref | 83/1493 | 5.3 | Ref |
| British Columbia | 44/495 | 8.2 | 1.08 (0.77-1.51) | 23/516 | 4.2 | 0.80 (0.51-1.24) |
| Quebec | 65/922 | 6.6 | 0.93 (0.66-1.29) | 42/945 | 4.2 | 0.85 (0.56-1.27) |
| Prairie provinces | 63/674 | 8.5 | 1.09 (0.79-1.49) | 39/697 | 5.2 | 0.98 (0.66-1.45) |
| Atlantic provinces | 23/278 | 7.6 | 1.11 (0.68-1.76) | 15/287 | 5.0 | 0.99 (0.53-1.74) |

\*Indigenous populations including First Nations, Inuit, or Métis.
Bolded values indicate significance at 95% confidence interval. Adjustment was for the other variables in the table. The number reporting COVID symptoms was 341 in unweighted analyses and 334 after survey weights were applied, showing the sampling frame as robust.

Table 3 shows the adjusted odds ratios of respondents or a member of the household being tested or scheduled for SARS-CoV-2 testing. The strongest determinant of testing was having COVID symptoms among members of the household, of whom about 16.5% were tested, compared to 2.1% among those without COVID symptoms among household members (OR=6.63, 4.46-9.79). Testing rates fell with age, but not significantly so. Testing rates were 2.7% in English and other European ethnicity, and higher in indigenous people (7.2%; OR=2.07; 1.13-3.64). However, the ORs fell notably (from 2.94 to 2.07) after adjustment for co-variates, suggesting residual confounding (data not shown). Testing rates in Quebec were roughly double those in Ontario, the reference province (OR=2.41; 1.49-3.96). Findings were similar among those who reported having been tested or being scheduled to be tested (data not shown).

**Table 3.** Logistic regression models for respondent or a member of household having been tested for SARS-CoV-2.

| Characteristics | Respondents or a member of household having been tested |  |  | Respondents only having been tested |  |  |
| --- | --- | --- | --- | --- | --- | --- |
|  | SARS-CoV-2 tested/not tested |  | Adjusted odds ratios (95% CI) | SARS-CoV-2 tested/not tested |  | Adjusted odds ratios (95% CI) |
|  | N=126/4015 | % |  | N=66/4075 | % |  |
| Respondent and/or household member had COVID symptoms |  |  |  |  |  |  |
| No | 80/3737 | 2.1 | Ref | 42/3775 | 1.1 | Ref |
| Yes | 46/278 | 16.5 | 6.63 (4.46-9.79) | 24/300 | 7.5 | 6.63 (3.82-11.32) |
| Gender |  |  |  |  |  |  |
| Male | 68/1921 | 3.5 | Ref | 34/1955 | 1.7 | Ref |
| Female | 58/2094 | 2.7 | 0.72 (0.51-1.03) | 32/2120 | 1.5 | 0.85 (0.52-1.38) |
| Age group |  |  |  |  |  |  |
| 18-45 years | 84/1858 | 4.5 | Ref | 41/1900 | 2.1 | Ref |
| 46-65 years | 32/1455 | 2.2 | 0.69 (0.45-1.05) | 19/1468 | 1.3 | 0.75 (0.42-1.31) |
| 66+ years | 10/702 | 0.7 | 0.54 (0.25-1.05) | 5/707 | 0.7 | 0.44 (0.14-1.08) |
| Education |  |  |  |  |  |  |
| Some college/university and higher | 96/3015 | 3.1 | Ref | 52/3059 | 1.7 | Ref |
| High school and under | 29/1000 | 2.8 | 0.88 (0.56-1.35) | 14/1016 | 1.3 | 0.74 (0.37-1.35) |
| Visible minority |  |  |  |  |  |  |
| No | 96/3519 | 2.7 | Ref | 52/3563 | 1.4 | Ref |
| Yes | 30/496 | 5.7 | 0.83 (0.36-1.05) | 14/512 | 2.7 | 0.54 (0.27-1.15) |
| Number of household members |  |  |  |  |  |  |
| Two people | 38/1548 | 2.4 | Ref | 19/1567 | 1.2 | Ref |
| Lived alone | 13/642 | 2.0 | 0.83 (0.43-1.52) | 13/642 | 2.0 | 1.77 (0.86-3.57) |
| Three people | 28/754 | 3.6 | 1.18 (0.69-1.97) | 18/764 | 2.3 | 1.28 (0.62-2.56) |
| Four or more people | 47/1071 | 4.2 | 1.32 (0.84-2.09) | 16/1102 | 1.4 | 0.93 (0.47-1.84) |
| Ethnicity |  |  |  |  |  |  |
| English and other European | 95/3462 | 2.7 | Ref | 54/3503 | 1.5 | Ref |
| Indigenous* | 16/206 | 7.2 | 2.07 (1.13-3.64) | 9/213 | 3.9 | 2.04 (0.89-4.28) |
| Others | 15/347 | 4.1 | 0.99 (0.49-1.89) | 4/359 | 1.0 | 0.47 (0.13-1.32) |
| Province/region |  |  |  |  |  |  |
| Ontario | 40/1537 | 2.5 | Ref | 22/1554 | 1.4 | Ref |
| British Columbia | 15/524 | 2.8 | 1.08 (0.59-1.93) | 7/532 | 1.3 | 0.81 (0.34-1.81) |
| Quebec | 47/940 | 4.8 | 2.41 (1.49-3.96) | 26/961 | 2.6 | 1.98 (1.05-3.81) |
| Prairie provinces | 19/717 | 2.6 | 1.13 (0.65-1.95) | 7/730 | 1.0 | 0.74 (0.32-1.63) |
| Atlantic provinces | 5/297 | 1.7 | 0.92 (0.34-2.14) | 4/299 | 1.2 | 1.03 (0.29-2.87) |

\*Indigenous populations including First Nations, Inuit, or Métis.
Bolded values indicate significance at 95% confidence interval. Adjustment was for the other variables in the table.

## Discussion

A nationally representative survey of Canadians finds that about 8% of adults report that they or someone in their household reported symptoms suggestive of COVID in March 2020. Being a visible minority was associated with higher self-reported COVID symptoms. Self-reported symptoms were notably less common at older ages than in younger adults. Only 3% of Canadian adults reported that they or someone in the household had been tested for SARS-CoV-2. The main determinants of being tested were the presence of COVID symptoms, being of Indigenous identity, and living in Quebec. Testing rates were somewhat lower in older adults. There are surprisingly few nationally representative studies, and despite some limitations, this study represents the first to document self-reported symptoms in a reasonably representative sample.

COVID symptoms overlap with some other infections, notably seasonal influenza, which could have inflated the coronavirus rates in this survey. However, a study comparing the COVID symptom syndrome with other infections in the United States suggests that most are actually due to COVID (Centers for Disease Control and Prevention, 2020). We found possible COVID symptoms to be less prevalent and the levels of testing marginally less prevalent in older adults than other age groups, despite the certainty that the vast majority of COVID hospitalizations and deaths occur at older ages. However, a weakness of our sample is the lack of representation from nursing and long-term care residents, in whom more than three-quarters of all COVID deaths occur (Eggertson and Wolfville, 2020). There may be additional reasons for the age-specific findings, however. Anecdotal reports suggest that older adults do not experience the symptoms used to define COVID infection in the poll, but may report vaguer symptoms such as dizziness and confusion (Graham, 2020). Further surveys focused on syndromes that might occur in older adults are warranted. The testing results are broadly consistent with reports of the general levels of access to SARS-CoV-2 testing during the survey time period, including a higher level of testing in Quebec than in Ontario (Angus Reid Institute, 2020).

This syndromic survey provides some insights into the prevalence of actual SARS-CoV-2 in Canada. The survey period was mostly in March, whereas models and case reports suggest that peak exposure and incidence of COVID occurred in mid-to late April (Eggertson and Wolfville, 2020). Hence, we would expect that a repeated syndromic survey should report a higher prevalence in April. On the other hand, Forum Research and Mainstreet Research reported an estimated prevalence of 5-8% in early-to-mid April in Ontario. However, they used a narrow range of symptoms from those we defined in our study (Forum Research and Mainstreet Research, 2020).

In the UK, pilot results from the COVID-19 Infection Survey being carried out by the Office for National Statistics found that 0.25% of the community population in England above the age of 2, tested positive for the SARS-CoV-2 antigen in early-to-mid May 2020. Testing involved home self-tests of nasal swabs, and excluded those in hospitals, care homes, or other institutional settings (Office for National Statistics, 2020). The prevalence was lower than expected in the pilot, and larger studies are planned, along with antibody surveys. A nationally representative sero-epidemiological study is needed to establish the population distribution of cumulative SARS-CoV-2 infection, which would capture both symptomatic and asymptomatic cases. We will soon conduct such a study in Canada called Action to Beat Coronavirus/Action pour Battre le Coronavirus (Ab-C). This study will determine the cumulative prevalence of SARS-CoV-2 infection during March-May 2020 in Canada through antibody testing, paired with household questionnaire data on COVID experience. A second round of questionnaires and antibody testing four to six months later in the same individuals will provide information on ongoing transmission, changes in the immune status of the population, and the durability of the immune response. The Angus Reid Forum is partnering with the Centre for Global Health Research at Unity Health Toronto and the University of Toronto on this study. The expected sample frame is 12-18,000 individuals surveyed, of which we expect 10-12,000 to agree to answer questions and provide a blood sample.

The present analysis informs the design of antibody surveys. For example, since the strongest determinant of COVID testing was the presence of symptoms, a similar higher prevalence of antibodies might be expected among those reporting symptoms (who may be more likely to enroll in the survey). Thus, ensuring that the sample size is sufficiently large to examine the seroprevalence in those without symptoms is key. Similarly, while the current study did not examine geographic clustering of COVID symptoms, this might well be the case with actual SARS-CoV-2 infection. This also necessitates a large and as diversely representative survey as possible. Older adults bear the brunt of COVID hospitalization and deaths. However, this population reported a lower prevalence of symptoms than younger adults. Thus, the Ab-C study will oversample the population age 60 or older, particularly to understand the possible role of asymptomatic infections at older ages. As the Ab-C study is not likely to capture the prevalence of infection among nursing home residents, ancillary studies are needed to quantify hazards among this important group.

### Limitations

Self-reported symptoms are, by their nature, subject to limitations and misclassification. However, the trends over time, even with misclassification, are useful for understanding trends in the actual underlying prevalence of infection. This is because the “noise” of COVID symptoms should change little from one survey to the next, and provided there is large “signal” due to the COVID pandemic—certainly the case with this virus—reliable estimation of the trends of infection are possible. Similar methodological insights have arisen from HIV-1 testing in pregnant populations in various countries (Kumar et al., 2006). The biases in enrollment are also inherent in any polling, but the Angus Reid polling showed reasonably good consistency with the Canadian 2016 census. Moreover, the testing results did not appear to be biased greatly, and the more common testing reported in Quebec is consistent with public health reports.

In conclusion, COVID surveillance using nationally representative surveys is essential, and ideally needs to be accompanied by antibody determination of infection. Particular attention must be paid to symptoms and testing levels in older adults.

## Materials and Methods

The Angus Reid Institute (ARI) conducted an online survey from April 1-5, 2020, among a representative randomized sample of 4,240 Canadian adults who are members of Angus Reid Forum, a national online sample of 50,000 Canadians used for political and other social polling (Angus Reid Institute, 2020). A probability sample of this size carries a margin of error of +/-2 percentage points, 19 times out of 20. The survey was commissioned and paid for by ARI. Questions were related to COVID and socio-demographic characteristics among both the respondents and members of their households (http://angusreid.org/covid-epidemiology-study/).

The outcomes were self-reported symptoms suggestive of COVID infection and testing for the virus. To understand the socio-demographic determinants of COVID symptoms, we conducted a logistic regression analysis, comparing COVID symptoms to the explanatory variables of gender, education level, province, age, ethnicity, visible minority (defined as persons, other than Aboriginal peoples, who are non-Caucasian in race or non-white in colour) (Statistics Canada, 2020b), and number of household members. We defined Indigenous as whether the person reported identifying with the Aboriginal peoples of Canada which includes First Nations, Métis or Inuit and/or those who reported Registered or Treaty Indian status (Statistics Canada, 2019). We also used logistic regression analysis to identify determinants of testing, which included the above explanatory variables and also COVID symptoms in the respondent or a family member. Respondents who did not report on any of the explanatory variables were excluded from these analyses. Discrepancies in or between totals are due to rounding. We applied to prevalences the survey weights, which are described earlier (Angus Reid Institute, 2020). We used RStudio Version 1.1.453 for analyses.

### Ethics approval

The Angus Reid Forum obtains consent from all participants, and the polling data without any individual identifiers are made available openly to bona-fide researchers. Ethics approval was not required as per Unity Health Toronto Research Ethics Board.

## Data Availability

Results of the Angus Reid poll are available through the link below

http://angusreid.org/wp-content/uploads/2020/04/2020.04.07_EpidemiologyCovidUpdate.pdf

